## Supplementary Table for "Characteristics of Long Covid: findings from a social media survey"

**Supplementary Table 1: Classification of ongoing symptoms by organ system**

| **Ongoing symptom** | **Organ system** |
| --- | --- |
| Abdominal Pain | GI |
| Diarrhoea | GI |
| Vomiting | GI |
| Nausea | GI |
| Loss of appetite | GI |
| Cough | Cardiopulmonary |
| Shortness of breath | Cardiopulmonary |
| Chest pain | Cardiopulmonary |
| Chest pressure | Cardiopulmonary |
| Chest tightness | Cardiopulmonary |
| Palpitations | Cardiopulmonary |
| Confusion | Neuro |
| Brain fog | Neuro |
| Poor concentration | Neuro |
| Depression | Neuro |
| Anxiety | Neuro |
| Memory problems | Neuro |
| Altered or loss of sense of smell | Neuro |
| Altered or loss of sense of taste | Neuro |
| Pins and needles | Neuro |
| Dizziness | Neuro |
| Tinnitus | Neuro |
| Exhaustion | Systemic |
| Sleep disturbance | Systemic |
| Fever | Systemic |
| Chills | Systemic |
| Sore throat | Nose/throat |
| Hoarse voice | Nose/throat |
| Sneezing | Nose/throat |
| Nasal symptoms | Nose/throat |
| Headache | Pain |
| Joint pain | Pain |
| Leg pain | Pain |
| Muscle aches | Pain |
| Skin rash | Skin |

**Supplementary Table 2: Pre-existing conditions in survey participants**

|  | Full sample | | Tested positive | | Tested negative or not tested | |
| --- | --- | --- | --- | --- | --- | --- |
|  | n | % | n | % | n | % |
|  | 2550 |  | 675 |  | 1793 |  |
| Allergies | 86 | 3.4 | 15 | 2.2 | 70 | 3.9 |
| Arthritis | 92 | 3.6 | 33 | 4.9 | 57 | 3.2 |
| Asthma | 344 | 13.6 | 99 | 14.7 | 240 | 13.5 |
| Cancer | 18 | 0.7 | 6 | 0.9 | 12 | 0.7 |
| Coeliac disease | 20 | 0.8 | 5 | 0.7 | 14 | 0.8 |
| Type 1 diabetes | 11 | 0.4 | 5 | 0.7 | 6 | 0.3 |
| Type 2 diabetes | 51 | 2.0 | 16 | 2.4 | 33 | 1.9 |
| Depression | 68 | 2.7 | 25 | 3.7 | 41 | 2.3 |
| Endometriosis | 29 | 1.2 | 5 | 0.7 | 24 | 1.3 |
| Epilepsy | 12 | 0.5 | 1 | 0.2 | 10 | 0.6 |
| Fibromyalgia | 61 | 2.4 | 18 | 2.7 | 41 | 2.3 |
| Hypertension | 141 | 5.6 | 42 | 6.3 | 88 | 4.9 |
| Hypothyroidism | 144 | 5.7 | 42 | 6.3 | 98 | 5.5 |
| Irritable bowel syndrome | 56 | 2.2 | 13 | 1.9 | 42 | 2.4 |
| Kidney disease | 12 | 0.5 | 2 | 0.3 | 10 | 0.6 |
| Liver disease | 18 | 0.7 | 4 | 0.6 | 14 | 0.8 |
| Migraine | 69 | 2.7 | 13 | 1.9 | 56 | 3.1 |
| Overweight/obesity | 38 | 1.5 | 14 | 2.1 | 24 | 1.3 |
| Polycystic ovary syndrome | 24 | 1.0 | 10 | 1.5 | 13 | 0.7 |
| Sleep disorders | 15 | 0.6 | 4 | 0.6 | 11 | 0.6 |

**Supplementary Table 3: Symptoms categorised by phase of illness**

|  | Did not experience | | Initial only (experienced during the two weeks of the illness) | | New symptom developed after acute phase (first two weeks of the illness) | | Initial symptom that persisted throughout the illness | |
| --- | --- | --- | --- | --- | --- | --- | --- | --- |
|  | n | % | n | % | n | % | n | % |
| Fever | 1206 | 47.7 | 1103 | 43.7 | 29 | 1.2 | 188 | 7.4 |
| Cough | 955 | 37.8 | 984 | 39.0 | 90 | 3.6 | 497 | 19.7 |
| Altered or loss of sense of smell | 1534 | 60.7 | 634 | 25.1 | 74 | 2.9 | 284 | 11.2 |
| Altered or loss of sense of taste | 1545 | 61.2 | 668 | 26.4 | 63 | 2.5 | 250 | 9.7 |
| Abdominal pain | 1751 | 69.3 | 348 | 13.8 | 215 | 8.5 | 212 | 8.4 |
| Diarrhoea | 1527 | 60.5 | 601 | 23.8 | 151 | 6.0 | 247 | 9.8 |
| Loss of appetite | 1483 | 58.7 | 760 | 30.1 | 101 | 4.0 | 182 | 7.2 |
| Nausea | 1700 | 67.3 | 414 | 16.4 | 189 | 7.5 | 223 | 8.8 |
| Vomiting | 2349 | 93.0 | 131 | 5.2 | 29 | 1.2 | 17 | 0.7 |
| Cognitive dysfunction* | 548 | 21.7 | 231 | 9.1 | 815 | 32.3 | 932 | 36.9 |
| Brain fog | 819 | 32.4 | 217 | 8.6 | 912 | 36.1 | 578 | 22.9 |
| Confusion | 1713 | 67.8 | 293 | 11.6 | 278 | 11.0 | 242 | 9.6 |
| Memory problems | 1277 | 50.6 | 155 | 6.1 | 776 | 30.7 | 318 | 12.6 |
| Poor concentration | 1105 | 43.8 | 283 | 11.2 | 693 | 27.4 | 445 | 17.6 |
| Depression | 2035 | 80.6 | 94 | 3.7 | 304 | 12.0 | 93 | 3.7 |
| Chest pain | 1186 | 47.0 | 449 | 17.8 | 352 | 13.9 | 539 | 21.3 |
| Chest pressure or tightness* | 586 | 23.2 | 611 | 24.2 | 307 | 12.2 | 1022 | 40.5 |
| Chest pressure | 927 | 36.7 | 629 | 24.9 | 290 | 11.5 | 680 | 26.9 |
| Chest tightness | 884 | 35.0 | 619 | 24.5 | 267 | 10.6 | 756 | 29.9 |
| Palpitations | 1191 | 47.2 | 273 | 10.8 | 585 | 23.2 | 477 | 18.9 |
| Shortness of breath | 640 | 25.3 | 516 | 20.4 | 328 | 13.0 | 1042 | 41.3 |
| Chills | 1154 | 45.7 | 999 | 39.6 | 83 | 3.3 | 290 | 11.5 |
| Dizziness | 1061 | 42.0 | 485 | 19.2 | 393 | 15.6 | 587 | 23.2 |
| Exhaustion | 271 | 10.7 | 421 | 16.7 | 340 | 13.5 | 1494 | 59.1 |
| Headache | 660 | 26.1 | 705 | 27.9 | 213 | 8.4 | 948 | 37.5 |
| Hoarse voice | 1706 | 67.5 | 367 | 14.5 | 171 | 6.8 | 282 | 11.2 |
| Nasal symptoms | 1614 | 63.9 | 441 | 17.5 | 196 | 7.8 | 275 | 10.9 |
| Sore throat | 1226 | 48.5 | 709 | 28.1 | 147 | 5.8 | 444 | 17.6 |
| Sneezing | 2180 | 86.3 | 158 | 6.3 | 107 | 4.2 | 81 | 3.2 |
| Tinnitus | 1791 | 70.9 | 73 | 2.9 | 396 | 15.7 | 266 | 10.5 |
| Joint pain | 1208 | 47.8 | 368 | 14.6 | 431 | 17.1 | 519 | 20.6 |
| Leg pain | 1585 | 62.8 | 273 | 10.8 | 371 | 14.7 | 297 | 11.8 |
| Muscle aches | 811 | 32.1 | 589 | 23.3 | 319 | 12.6 | 807 | 32.0 |
| Pins and needles | 1688 | 66.8 | 171 | 6.8 | 451 | 17.9 | 216 | 8.6 |
| Skin rash | 2040 | 80.8 | 187 | 7.4 | 199 | 7.9 | 100 | 4.0 |
| Sleep disturbance | 1160 | 45.9 | 414 | 16.4 | 463 | 18.3 | 489 | 19.4 |
| Other |  |  |  |  |  |  |  |  |
| Eye | 2401 | 95.1 | 44 | 1.7 | 59 | 2.3 | 22 | 0.9 |
| Ear (excluding tinnitus) | 2399 | 95.0 | 35 | 1.4 | 74 | 2.9 | 18 | 0.7 |
| Tachycardia | 2476 | 98.0 | 6 | 0.2 | 40 | 1.6 | 4 | 0.2 |
| Hair loss | 2474 | 97.9 | 14 | 0.6 | 35 | 1.4 | 3 | 0.1 |
| Acid reflux | 2488 | 98.5 | 9 | 0.4 | 26 | 1.0 | 3 | 0.1 |

*Cognitive dysfunction and chest pressure or tightness are combined derived variables made up of component symptoms. The combined derived variable does not directly reflect the pattern of the component symptoms at each phase as the overall distribution is considered for the combined derived variable. For example, brain fog exhibited as a new symptom after the acute phase in 36.1% but cognitive dysfunction at this phase was 32.3% as a proportion of those reporting brain fog in this phase had reported experiencing confusion, memory problems and/or poor concentration as start and/or ongoing symptoms which was taken into account for the derivation of the combined variable.

**Supplementary Table 4: Duration and pattern of illness in those who reported full recovery from Long Covid**

|  | Recovered | | | | | |
| --- | --- | --- | --- | --- | --- | --- |
|  | Full sample | | Tested positive | | Test negative/not tested | |
|  | n | % | n | % | n | % |
|  | 58 |  | 14 |  | 41 |  |
| Duration of illness |  |  |  |  |  |  |
| 1-2 months | 17 | 29.3 | 5 | 35.7 | 11 | 26.8 |
| 2-3 months | 11 | 19.0 | 1 | 7.1 | 10 | 24.4 |
| 3-4 months | 10 | 17.2 | 5 | 35.7 | 4 | 9.8 |
| 4-5 months | 5 | 8.6 | - | - | 5 | 12.2 |
| 5-6 months | 7 | 12.1 | - | - | 7 | 17.1 |
| 6-9 months | 8 | 13.8 | 3 | 21.4 | 4 | 9.8 |
| Time since last symptom |  |  |  |  |  |  |
| ≤2 weeks | 3 | 5.2 | 2 | 14.3 | 1 | 2.4 |
| >2weeks - <1 month | 8 | 13.8 | 1 | 7.1 | 6 | 14.6 |
| 1-2 months | 12 | 20.7 | 3 | 21.4 | 9 | 22.0 |
| 2-3 months | 12 | 20.7 | 4 | 28.6 | 7 | 17.1 |
| 3-4 months | 7 | 12.1 | 3 | 21.4 | 4 | 9.8 |
| >4 months | 16 | 27.6 | 1 | 7.1 | 14 | 34.2 |
| Pattern of symptoms in last month of illness |  |  |  |  |  |  |
| Gradually got better | 25 | 43.1 | 6 | 42.9 | 17 | 41.5 |
| Fluctuating | 9 | 15.5 | 2 | 14.3 | 6 | 14.6 |
| Come and go | 24 | 41.4 | 6 | 42.9 | 18 | 43.9 |
