## Supplementary Figure for "Characteristics of Long Covid: findings from a social media survey"

**Supplementary Figure 1: Reported SARS-CoV-2 testing history in survey participants**

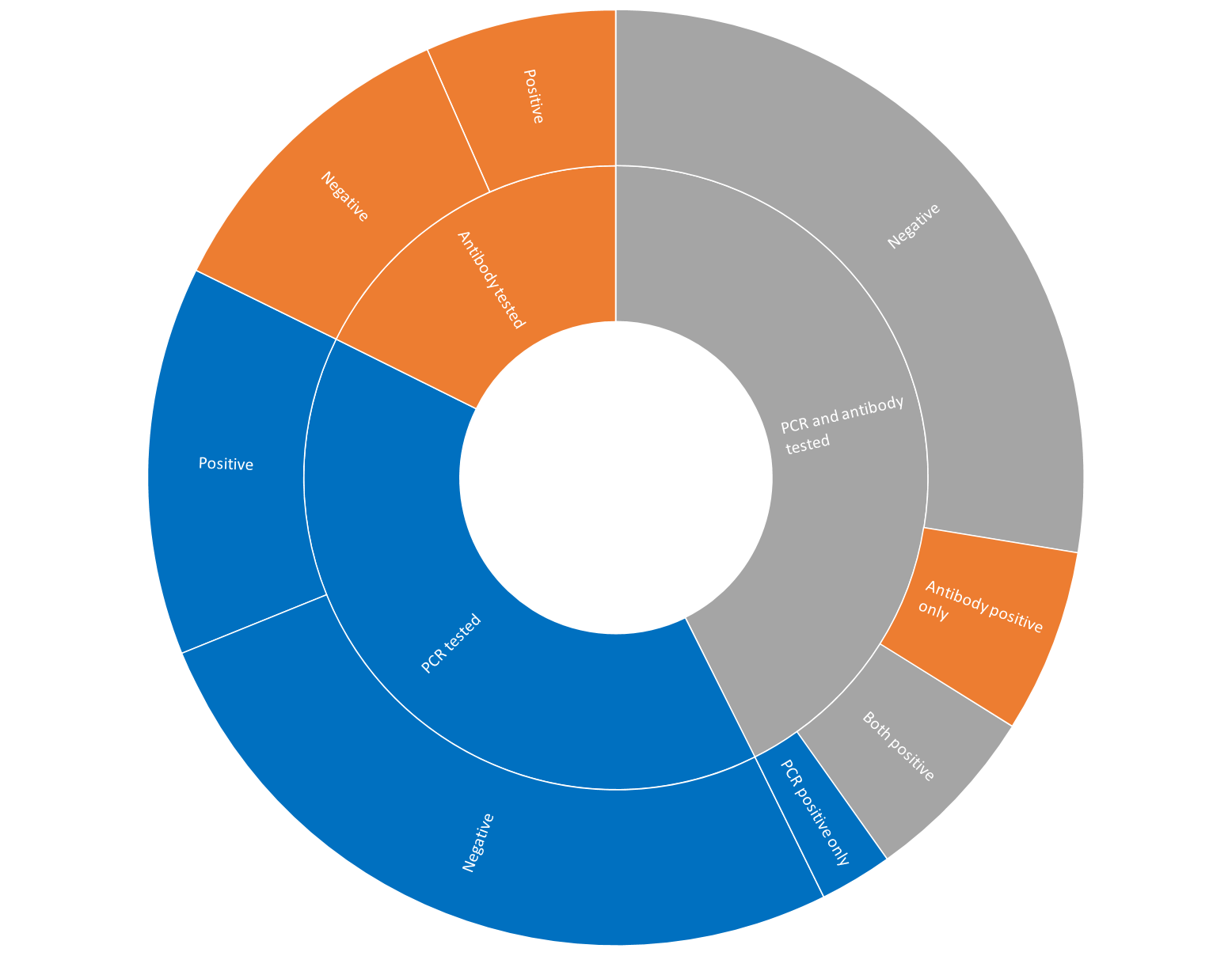

**Supplementary Figure 2: Silhouette coefficient for 2 to 10 clusters**

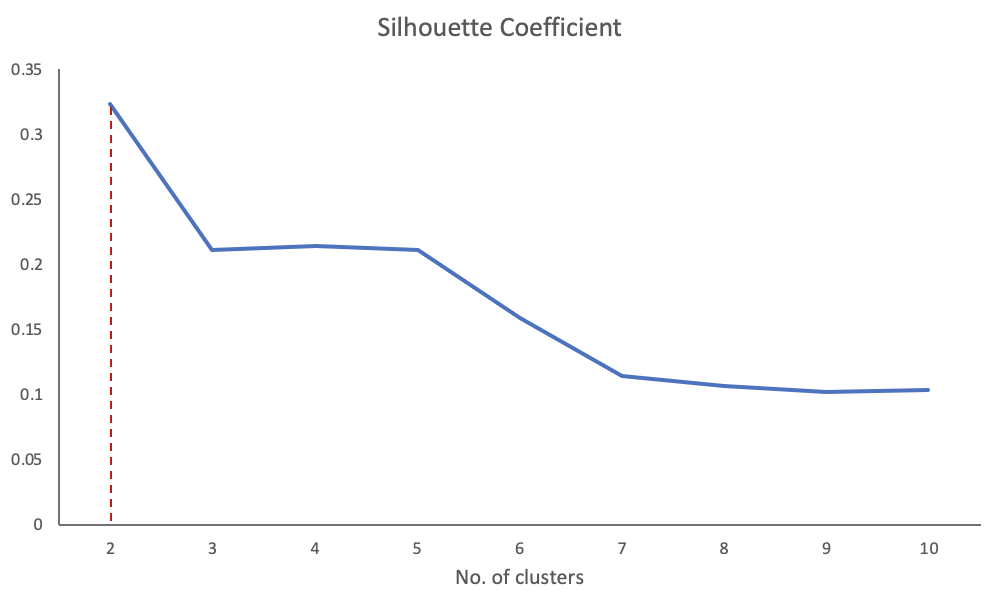

**Supplementary Figure 3: Two clusters of acute symptoms and ongoing symptoms among these clusters**

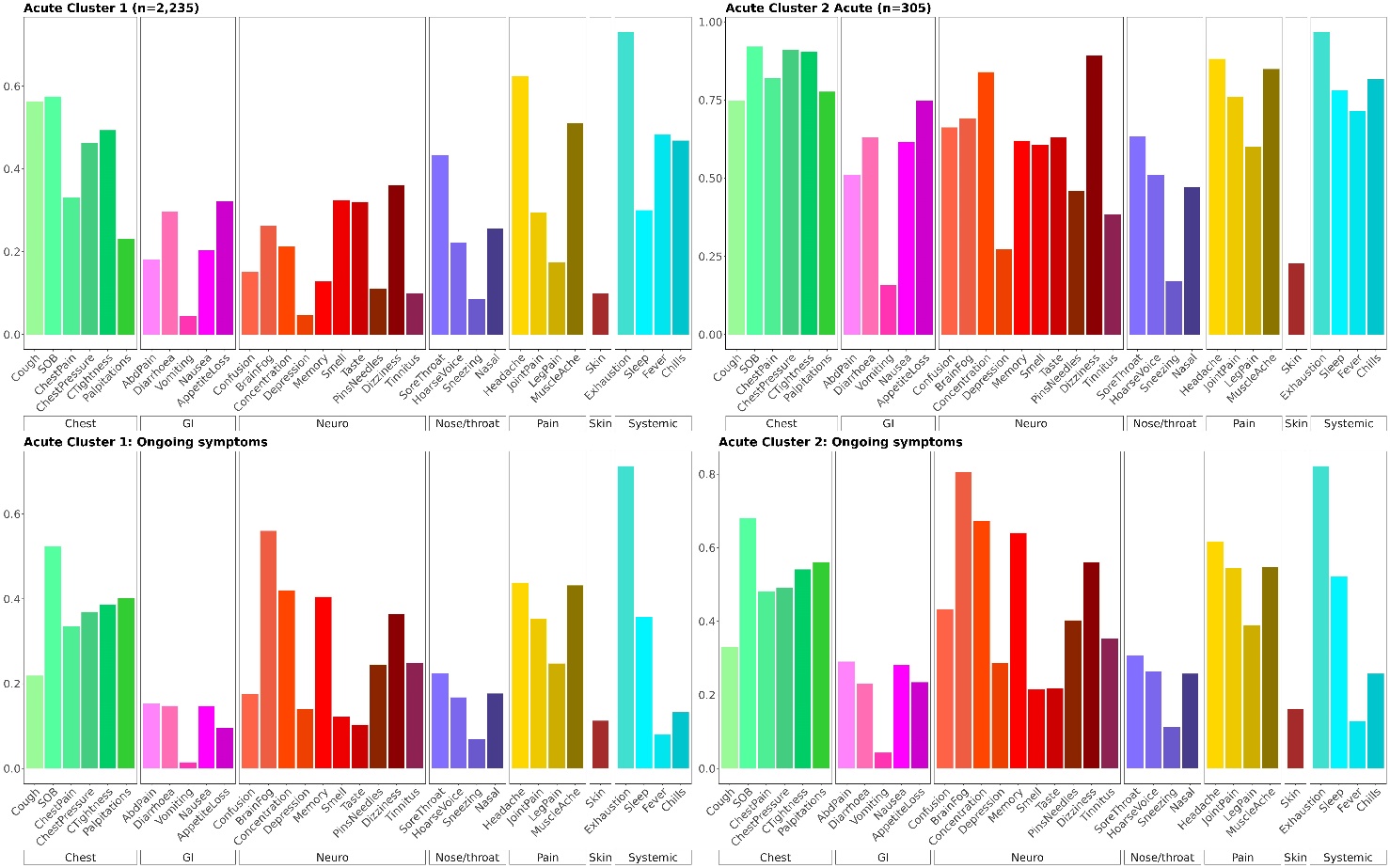

**Supplementary Figure 4: Clustering of lab confirmed subgroup only identifies similar clusters to whole dataset**

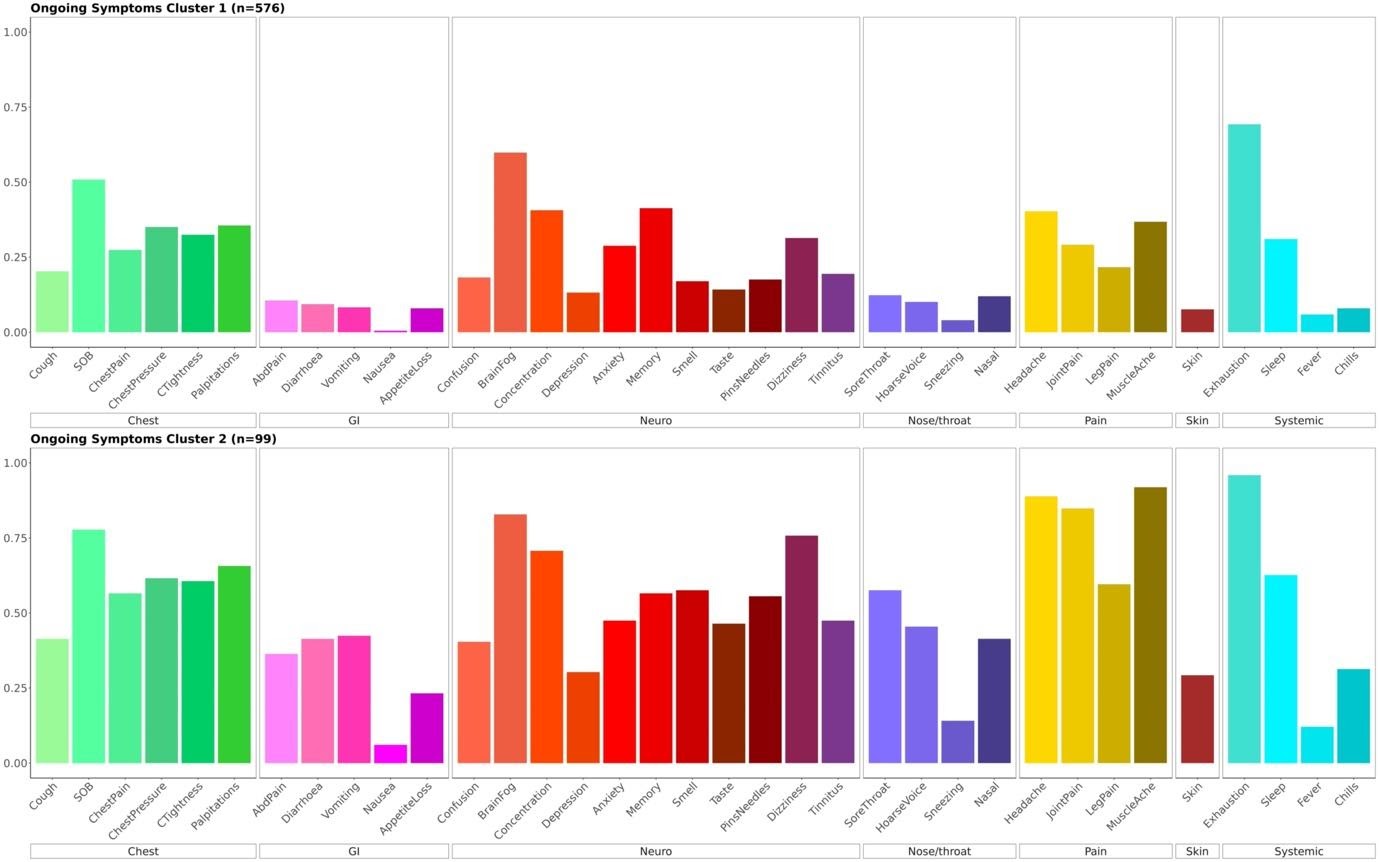

**Supplementary Figure 5: Transition from acute symptom clusters to ongoing symptom clusters by number of affected systems**

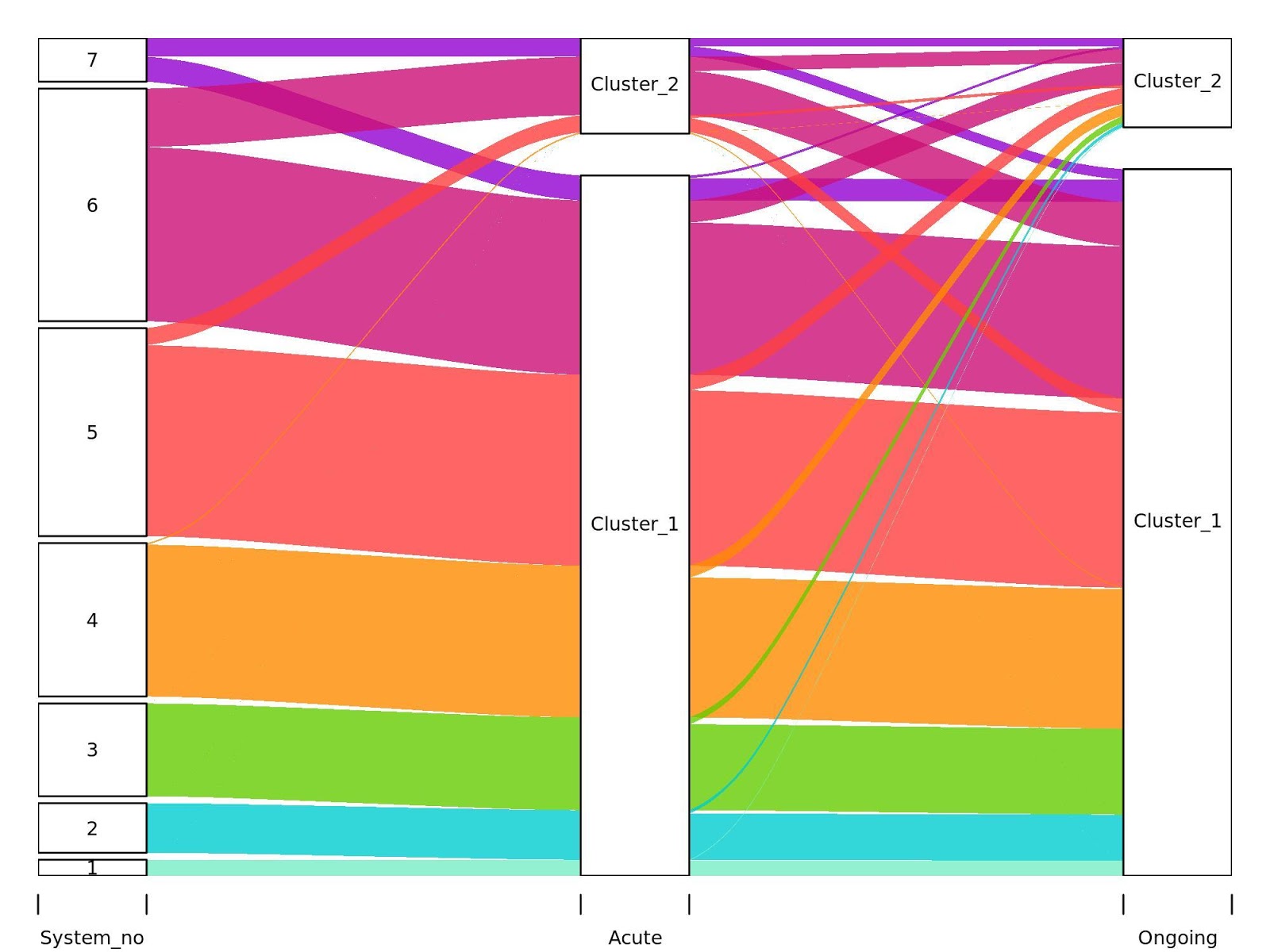

**Supplementary Figure 6: Mutually adjusted predictors of transition from acute symptom cluster 1 (ASC1: cardiopulmonary predominant) to ongoing symptom cluster 2 (OSC2: multisystem)**

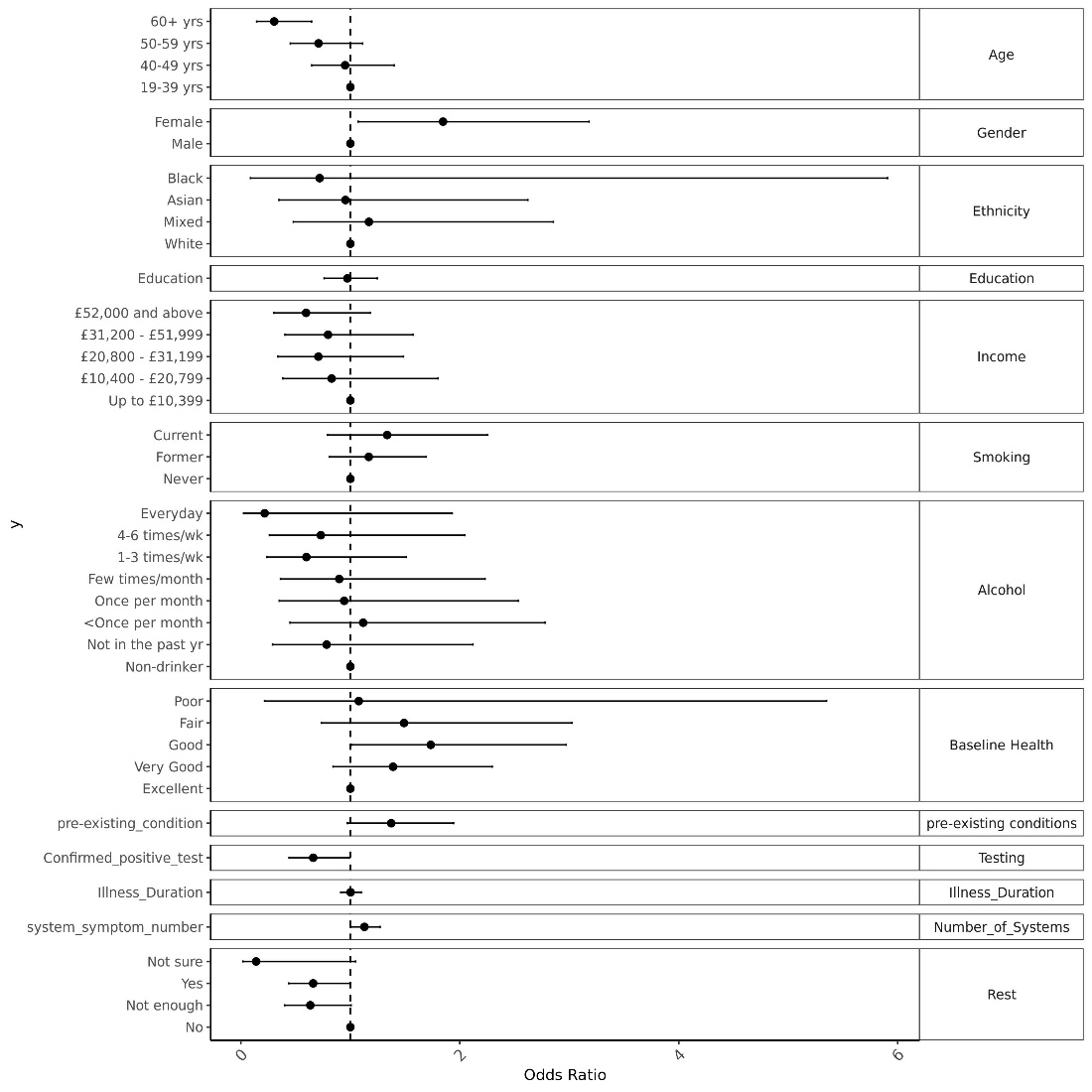
